# Sequencing SARS-CoV-2 from Antigen Tests

**DOI:** 10.1101/2021.07.14.21260291

**Authors:** Ashley Nazario-Toole, Holly M. Nguyen, Hui Xia, Dianne N. Frankel, John W. Kieffer, Thomas F. Gibbons

## Abstract

Genomic surveillance empowers agile responses to SARS-CoV-2 by enabling scientists and public health analysts to issue recommendations aimed at slowing transmission, prioritizing contact tracing, and building a robust genomic sequencing surveillance strategy. Since the start of the pandemic, real time RT-PCR diagnostic testing from upper respiratory specimens, such as nasopharyngeal (NP) swabs, has been the standard. Moreover, respiratory samples in viral transport media are the ideal specimen for SARS-CoV-2 whole-genome sequencing (WGS). In early 2021, many clinicians transitioned to antigen-based SARS-CoV-2 detection tests, which use anterior nasal swabs for SARS-CoV-2 antigen detection. Despite this shift in testing methods, the need for whole-genome sequence surveillance remains. Thus, we developed a workflow for whole-genome sequencing with antigen test-derived swabs as an input rather than nasopharyngeal swabs. In this study, we use excess clinical specimens processed using the BinaxNOW™ COVID-19 Ag Card. We demonstrate that whole-genome sequencing from antigen tests is feasible and yields similar results from RT-PCR-based assays utilizing a swab in viral transport media.

## Introduction

Early in the pandemic Mercatelli and Giorgi predicted a low mutation rate for SARS-CoV-2 based on whole-genome sequencing of 48,635 specimens [1]. Continued genomic surveillance has revealed a worrying mutation rate of 3.7×10^−6^ per nucleotide per cycle [2]. Furthermore, the rate of mutation varies across the SARS-CoV-2 genome, with several sites exhibiting recurrent mutations that, due to strong positive selection, emerged independently a minimum of three times in strains sequenced across the globe [3]. Ongoing genomic surveillance of SARS-CoV-2 is critical for identifying emerging variants [4,5]; driving changes to public health policies [6]; and confirming cases of reinfection [7].

### Variants

The CDC defines a variant as a viral genome that has one or more mutations that differentiate it from other variants in circulation [8]. Variants may have different potential impacts on public health. Variants of interest (VOIs) require monitoring. Variants of concern (VOCs) may affect treatment, transmission, or disease severity. Variants of high consequence (VOHC) have significantly reduced effectiveness of prevention measures or therapeutics relative to existing or previous variants. Fortunately, no VOHCs have been identified to date. The following VOCs highlight the challenges faced by clinicians and public health officials: B.1.1.7 (UK) increased transmission, P.1 (Brazilian) reduced serum neutralization, and B.1.351 (South African) reduced vaccine efficacy in clinical trials conducted by Pfizer and Novavax [9–12]. Increased transmission of regional strain variants may prompt enacting or increasing social distancing, mask wearing, and/or travel restriction.

### Reinfection

Although exceedingly rare, reinfection has been documented and is a threat to vulnerable populations. Although most suspected cases of reinfection were a resurgence of the same viral strain that initially infected the patient, other cases demonstrate reinfection with genetically distinct genomes [7,13]. Genome surveillance enables us to monitor for cases of reinfection and discern whether these cases are associated with particular variants.

### Specimen Sources

Initially, SARS-CoV-2 tests relied on nasopharyngeal swabs placed in viral transport media (VTM) followed by RT-PCR [14–16]. In May 2020, the FDA approved the first emergency use authorization for an antigen test of SARS-CoV-2 [17]. In early 2021, the DoD began using antigen testing on asymptomatic active duty and DoD civilian personnel [18,19]. Antigen tests meet the World Health Organization’s (WHO) definition of an Affordable, Selective and Sensitive, User-friendly, Rapid and Robust, Equipment-free, Deliverable to end-users (ASSURED) diagnostic tool [20,21]. Thus, antigen testing has been a suitable tool for global monitoring of new SARS-CoV-2 cases. Currently, most pipelines for WGS rely on upper respiratory tract clinical diagnostic samples with high viral loads [22,23]. However, antigen tests use anterior nasal (AN) swabs. NP and AN swabs differ in the location from which specimen is derived from the patient [24–26]. Additionally, NP swabs are stored in up to 3 milliliters (mL) of viral transport media or buffered saline solutions whereas AN swabs are inserted into an antigen card with only a few drops of proprietary extraction buffer [16,25]. Finally, after collection, samples in viral transport media are stored in freezers; where antigen cards may be stored at a range of temperatures. All of these pre-analytical factors present concerns regarding the quality of specimen to be used for whole-genome sequencing.

### Sequencing considerations

To continue whole-genome sequences at institutions that have adopted antigen testing, communication with clinicians is required to minimize pre-analytical factors that may lead to sample degradation or discarding of samples. Institutions that choose to adopt antigen-based whole genome sequence pipelines must work with clinicians and institutional review boards (IRBs) to ensure that clinical specimens are retained for genome surveillance purposes. Clinical researchers must also work with clinicians to ensure proper storage of antigen cards prior to transportation to the sequencing laboratory.

Here, we tested various collection and storage parameters to optimize sample preparation for SARS-CoV-2 whole genome sequencing from Ag-cards. We demonstrate that it is possible to produce high quality genomic surveillance data from antigen-derived AN swabs. Specifically, we validated PCR-based whole genome sequencing from AN swabs from the BinaxNOW™ COVID-19 Ag Card (Abbott Diagnostics Scarborough).

## Results

### Workflow

Our optimized workflow, shows how de-identified, excess clinical specimens of positive BinaxNOW™ COVID-19 Ag Cards are processed (**Figure 1**). Per CDC [18,27] and BMBL 6^th^ Edition [28] guidelines for handling SARS-CoV-2, the antigen cards are handled in a biosafety cabinet in a biosafety level 2 (BSL2) room. Flocked swabs are removed from the antigen card and placed into 500 µL of DNA/RNA Shield in a 15 mL conical tube. Swabs are then broken at the breakpoint, the tube cap securely fastened, and the sample vortexed vigorously for 15-30 seconds. Samples are stored at 4°C until RNA extraction. Viral load is determined using a modified version of the CDC research use only (ROU) real-time RT-PCR assay, which targets the viral nucleocapsid (CDC 2019-nCoV N1) and human RNase P (RP). Samples with an N1 cycle threshold (C_T_) less than 25 have sufficient quantity of SARS-CoV-2 RNA for sequencing library preparation (Paragon Genomics CleanPlex Flex SARS-CoV-2 Panel). Prepared libraries are checked using fragment analysis and a library quality ratio score is calculated as previously described [29]. Briefly, quality ratio scores are calculated by dividing the concentration [ng/µL] of target amplicons (fragments 250 base pairs (bp) to 350 bp) by the concentration of nonspecific bands (50 base pairs (bp) to 190 bps). Samples with a quality score greater than or equal to one are sequenced using Illumina’s MiSeq or NextSeq500 systems. Finally, sequence data is processed through a user-defined bioinformatics pipeline.

**Figure 1.**
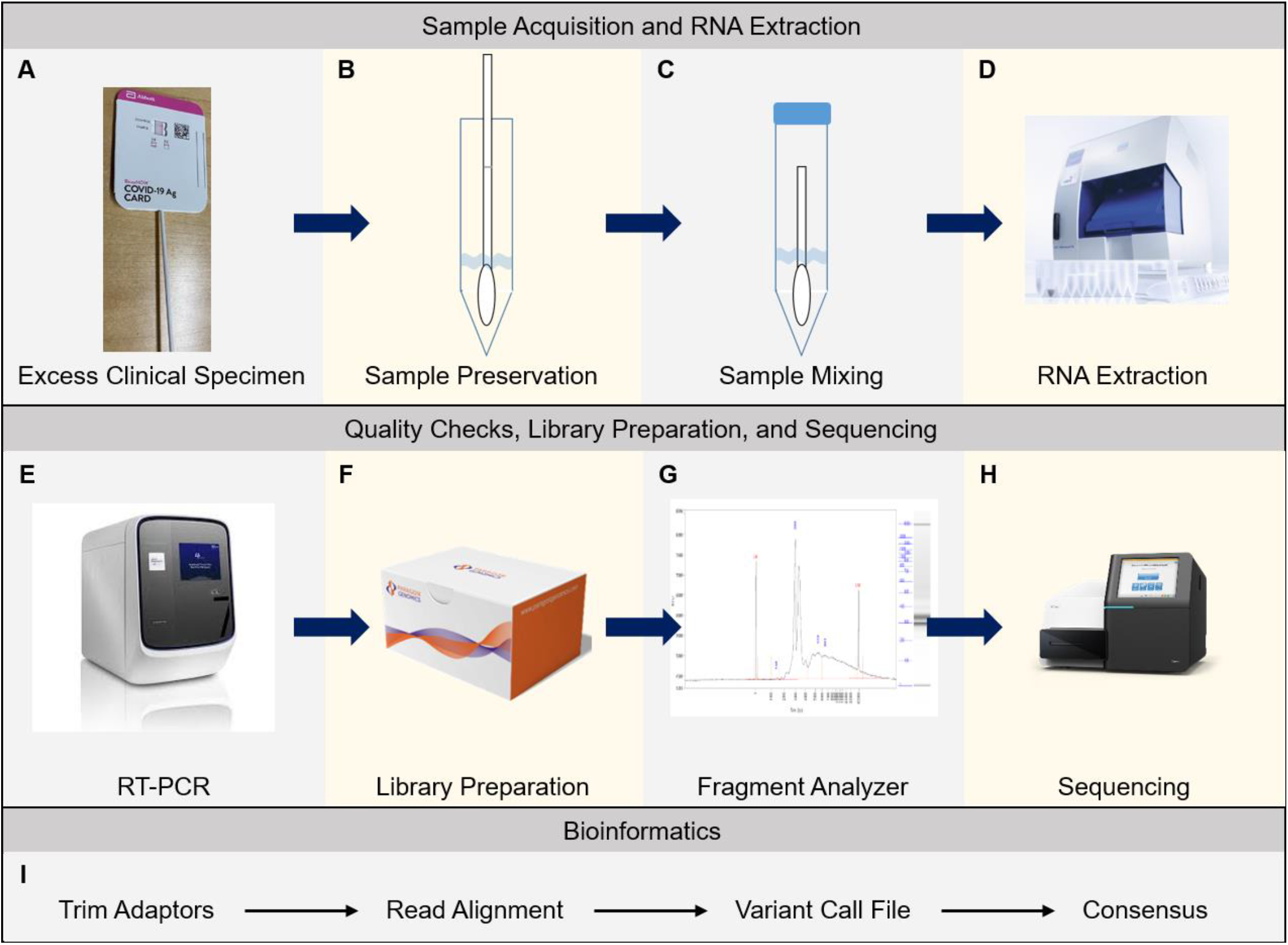
Workflow. (A) BinaxNOW™ antigen cards are run per manufacturer guidelines. (B-C) Swabs are removed, placed in 500 µL DNA/RNA shield, broken at the breakpoint, capped and vortexed in a biosafety cabinet (23). (D) RNA is extracted. (E) RT-PCR is used to measure viral load as N1 C_T_. (F) Library preparations are made for samples with C_T_ values less than 25. (G) Fragment analysis is used to check library quality and (H) samples with a quality score of equal to or greater than 1 are sequenced. (I) An in-house bioinformatics pipeline is used to process sequence data.

### Sample Preparation Optimization

To optimize the sample collection and preparation methods, we utilized a previously sequenced SARS-CoV-2 positive NP specimen with an N1 C_T_ of 12.28 (#5195) (**Fig 2**). First, we examined the potential for sample loss and degradation due to (1) exposure to the BinaxNOW™ proprietary extraction buffer and (2) storage conditions (**Fig 2A**). In this, and every following experiment, we included a standardized positive control, water extraction, and no template control. The positive control result is shown; however, the water extraction and template controls are not shown, due to their expected and observed lack of amplification. A BinaxNOW™ COVID-19 Ag Card swab was dipped into NP specimen # 5195 and then placed into 500 µL DNA/RNA shield (*swab, no Ag test*). For comparison, a second swab dipped into the same NP specimen, run through the antigen test for 15 min per manufacturer’s instructions, and then placed into DNA/RNA shield (*swab*). The N1 C_T_ of the “swab, no Ag test” sample was 15.75 while the N1 C_T_ of the “swab” sample was 15.82, indicating that little to no viral RNA was lost after 15 min exposure to the Ag-card extraction buffer. To test if prolonged storage of antigen cards might affect sample yield, a third swab was dipped into the NP specimen, run through the antigen test, and the whole card was stored at 4°C for 2 hours before the swab was placed in DNA/RNA Shield (*swab & storage*). Again, RT-PCR results revealed that there was no sample degradation, as observed through N1 C_T_ values of 15.82 (*swab*) and 15.60 (*swab & storage*). PCR-amplicon sequencing libraries were then prepared for each sample using the Paragon Genomics CleanPlex Flex SARS-CoV-2 Panel. For a library prep and sequencing control, we also prepared a library using commercially available SARS-CoV-2 genomic RNA (ATCC # VR-1986D, SARS-Cov-2 Isolate USA-WA1/2020). Library quality scores were calculated as previously described and samples were sequenced at 2×151 bp reads on the Illumina MiSeq system. N1 C_T_, library quality ratio scores, 20X genome coverage (20 or more reads mapped per nucleotide), and viral PANGO lineages are shown in **Figure 2B**. As expected, the positive control VR-1986D was assigned to PANGO lineage A and all dipped Ag-card specimens were assigned to PANGO lineage B.1, the same lineage assigned to NP specimen 5195. IGV snapshots show that all samples map at greater than 99.5% at 20X coverage across the genome (**Fig 2C**). Together these finding indicated that sample exposure to Ag-card buffer and testing conditions do not impact the quality of RNA and subsequent sequencing steps.

**Figure 2.**
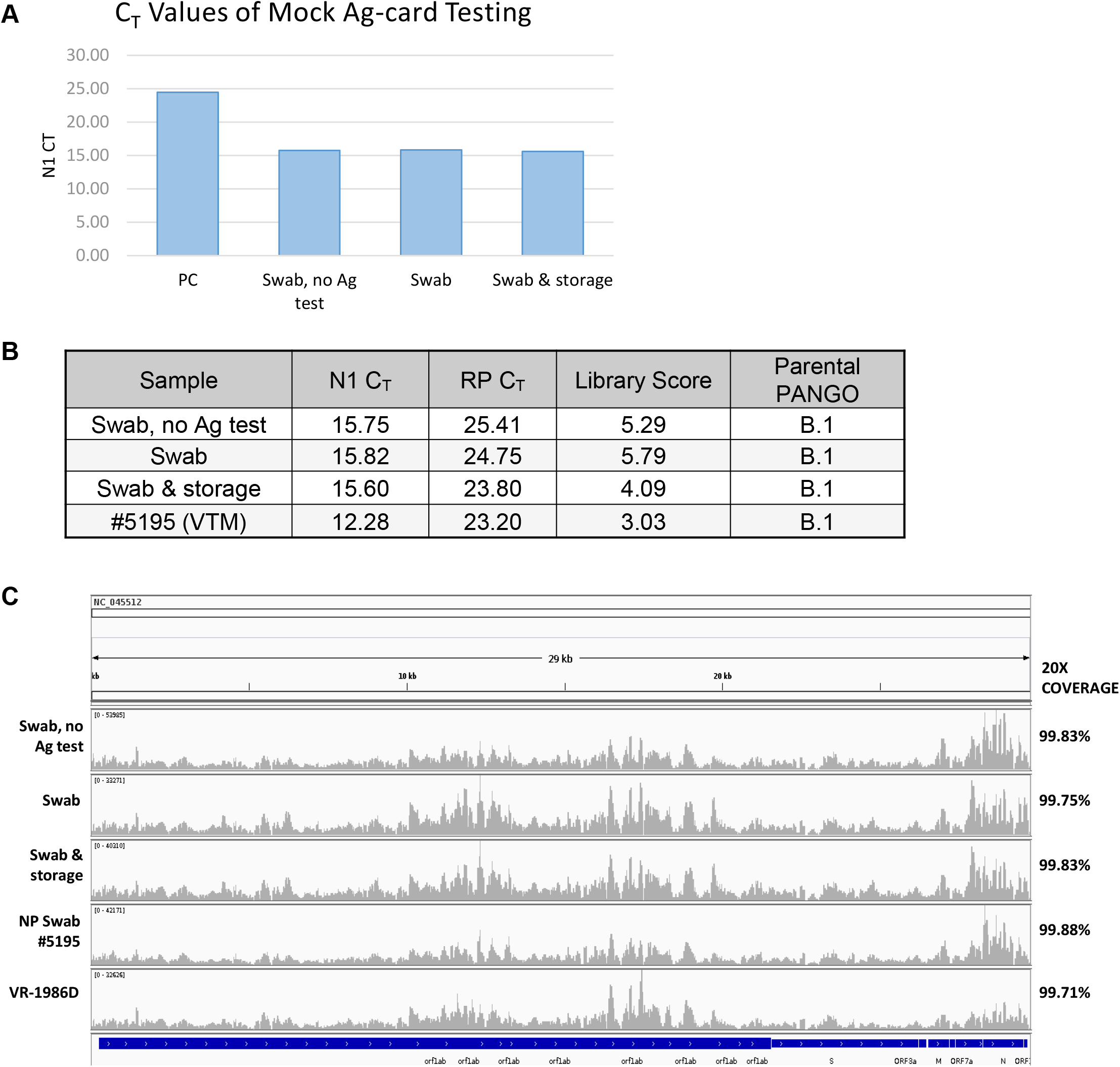
Assay Development. (A) An NP specimen was used to evaluate the feasibility of obtaining viral RNA from Ag-card derived specimens. RT-PCR was carried out on specimens collected under each condition. (B) RT-PCR N1 and RP Ct values, sequence library quality scores, and viral PANGO lineage assignments of antigen card specimens and reference NP specimen #5195. (C) IGV screen shots of sample SARS-CoV-2 genome coverage. VR-1986D = Positive Control Genomic RNA from SARS-Cov-2, Isolate USA-WA1/2020.

### Performance Comparison

Given that we carried out our preliminary tests in a mock fashion by dipping swabs in a previously tested NP specimen, we next utilized COVID-19 positive Ag-cards to test the viability of sequencing from real specimens (excess clinical specimens collected under IRB-approved protocol FWH20200103E). Seven completed SARS-CoV-2 positive BinaxNOW™ COVID-19 Ag Cards were delivered to the lab and immediately processed as follows: Antigen cards were disassembled and the AN swabs and positive line of the lateral flow strips were separately placed into DNA/RNA shield, stored at 4°C for 48 hours, extracted, then amplified using RT-PCR (**Fig 3A**). We hypothesized that due to the nature of the lateral flow assay, viral RNA would be concentrated on the positive line of the strip. We found that swabs slightly outperformed the positive line by yielding higher viral loads in all samples. Specifically, swab N1 C_T_ values were, on average, 1.57 C_T_s lower than the line (± 1.16 C_T_s) (p-value = 0.0116). Interestingly, host RNA levels were significantly higher in the swabs: RP was detected in swab specimens 4.43 cycles sooner than the line (± 1.30 C_T_s) (p-value = 0.0001). Thus, relative to host RNA, viral RNA is enriched on the positive line. As we were able to detect comparable levels of viral RNA on swabs and positive lines, we next prepared libraries from three samples with N1 C_T_ < 25 from both the swab and positive line (**Fig 3B** and **3C**). Regardless of the sample source, sequenced specimens had 20x genome coverage (20 or more reads per nucleotide) of greater than or equal to 99%, signifying that both the swab and positive line from antigen cards can be used for whole-genome sequencing with no loss in coverage.

**Figure 3.**
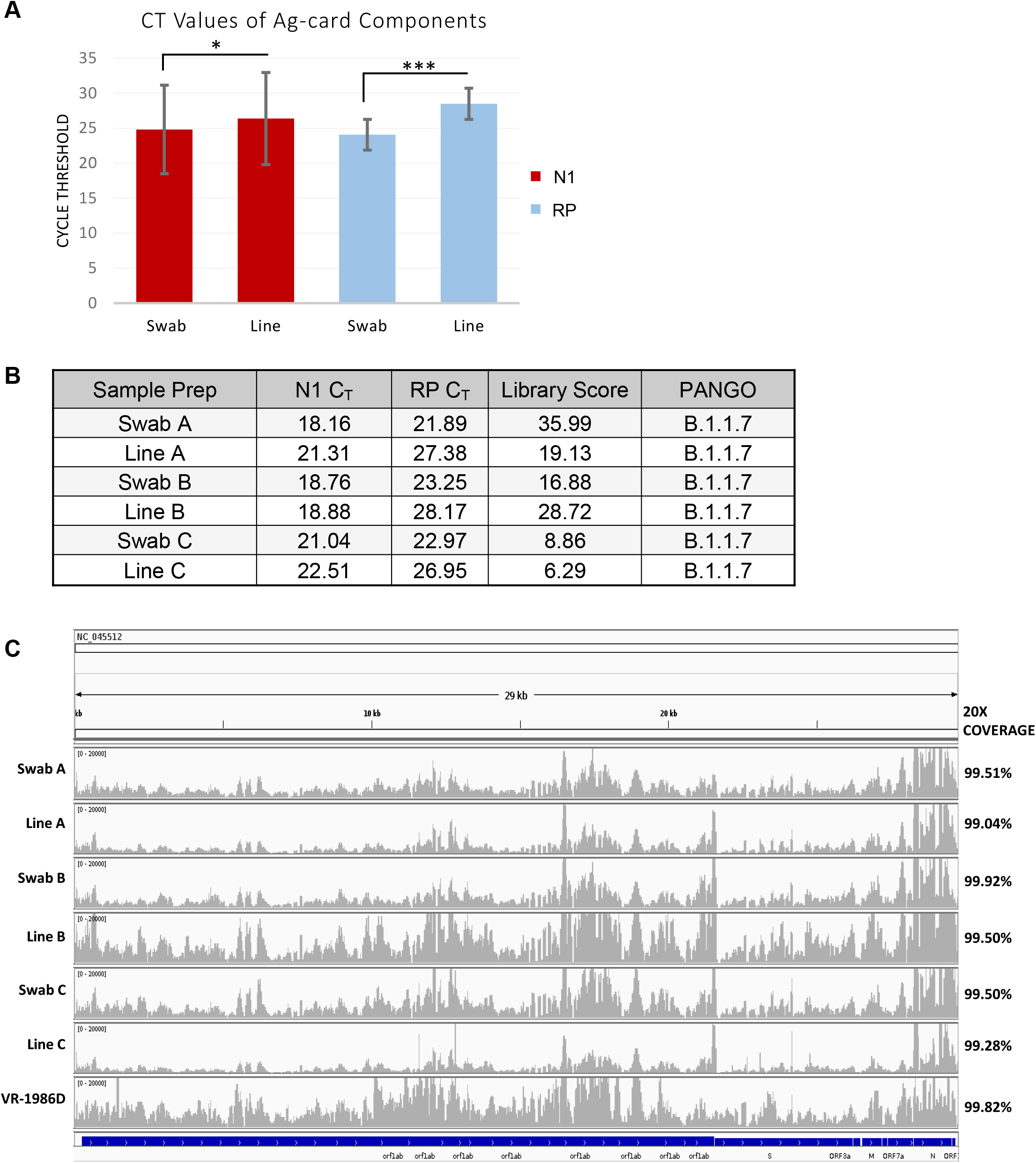
Sample source performance comparison. (A) SARS-CoV-2 positive BinaxNOW™ COVID-19 Ag Cards were used to evaluate which part of the card, swab or lateral flow strip positive line, yielded the highest quantity and quality of viral RNA. Extracted RNA was tested for N1 and RP levels using RT-PCR. The average N1 and RP C_T_ values are plotted. n = 7 cards. Statistical analysis = Two-tailed, paired *t-*test, * p<0.05; *** p<0.0005 (B) N1 and RP C_T_ values, library quality scores and PANGO lineage assignments. (C) IGV screen shots of SARS-CoV-2 genome coverage.

**Figure 3**. Sample source performance comparison. (A) SARS-CoV-2 positive BinaxNOW™ COVID-19 Ag Cards were used to evaluate which part of the card, swab or lateral flow positive line, yielded the highest quantity and quality of viral RNA. Extracted RNA was tested for N1 and RP levels using RT-PCR. The average N1 and RP C_T_ values are plotted. n = 7 cards. Statistical analysis = Two-tailed, paired *t-*test, * p<0.05; *** p<0.0005 (B) N1 and RP Ct values, library quality scores and PANGO lineage assignments. (C) IGV screen shots of SARS-CoV-2 genome coverage.

## Discussion

When the COVID-19 pandemic started, highly sensitive and specific real-time RT-PCR tests were the first to be developed, quickly becoming the mainstay of diagnostic testing [30,31]. However, RT-PCR tests require specialized kits, equipment, personnel, and laboratories. These factors lead to slower turnaround times as compared to recently released rapid diagnostic tests - which can be readily employed to screen populations such as basic military trainees. With more clinicians and point-of-care testing sites adopting antigen-based SARS-CoV-2 tests, fewer RT-PCR specimens may be available to support genomic surveillance of viral variants worldwide. To facilitate future genomic surveillance efforts, we developed a workflow for SARS-CoV-2 whole-genome sequencing from antigen-based test specimens (**Fig 1**). We were able to recover viral RNA of sufficient quantity and quality for targeted sequencing of the SARS-CoV-2 genome (**Fig 2**). We found that the quality of genome sequences derived from Ag-test samples is comparable to RNA isolated from NP swabs collected for RT-PCR (**Fig 2**). Furthermore, a comparison of sample sources, antigen card swab vs. lateral flow assay (LFA) positive line, showed that viral RNA from both sources can generate high quality sequencing libraries (**Fig 3**). For ease of use and biosafety concerns, we recommend collecting the swab over the LFA positive line as both specimen types produced comparable libraries.

Antigen card specimens present a few limitations when compared to NP swabs collected in viral transport media (VTM). First, in the workflow presented here, antigen test swabs are stored in 500 µL of DNA/RNA shield while NP swabs are stored in up to 3 mL of VTM. Multiple RNA extractions can be carried out from VTM specimens after the initial round RT-PCR testing but, antigen swab-derived viral RNA can only be extracted once. Next, variants with novel or interesting mutations can be cultured from VTM; enabling downstream biochemical or viral neutralization assays. In contrast, antigen tests immediately expose the specimen to an extraction buffer which disrupts the viral membrane, releases viral RNA, and enables the presentation of viral nucleocapsid antigens to anti-nucleocapsid antibodies on the LFA positive line. Thus, viruses collected post antigen-test are most likely non-culturable, however we did not evaluate cultivability.

Here we only attempted to sequence from the BinaxNOW™ antigen test, but other antigen tests may also yield viable specimens for whole-genome sequencing. This work is a first step for future studies examining the sequencing utility of specimens collected for other antigen or rapid molecular tests.

## Materials and Methods

### Specimen Acquisition

The BinaxNOW™ COVID-19 Ag Card (Abbott) was used to test basic medical trainees for SARS-CoV-2. Excess clinical specimens were de-identified, the entire card placed in a plastic biohazard bag, and samples transferred to the Clinical Investigations and Research Support (CIRS) laboratory. Excess clinical specimens were obtained under an institutional review board (IRB) exempt protocol (IRB reference number FWH20200103E).

### Specimen Preparation

The antigen card was disassembled in a biosafety cabinet. The swab was placed in a 15 mL conical tube with 500 µL DNA/RNA Shield (Zymo Research, Catalog #R1100) and stored at 4°C until the RNA was extracted. The positive line of the LFA test strip was excised and placed in 500 µL of DNA/RNA shield and stored in a 2 mL cryovial at 4°C until the RNA was extracted.

### RNA Extraction

Samples were extracted using the EZ1 Virus Mini Kit v2.0 (Qiagen, Catalog # 955134), per the manual. Briefly, 400 µL of sample was extracted and eluted into 60 µL AVE buffer (supplied in the kit). The following controls were run with each extraction: a positive control – a nasopharyngeal swab of SARS-CoV-2 diluted to achieve a C_T_ of approximately 25 - and a negative control consisting of 200 µL nuclease-free water and 200 µL of DNA/RNA Shield. The extracted RNA was frozen at -80°C or used immediately for RT-PCR.

### RT-PCR

The following primers were utilized: N1 Forward, 5’-GACCCCAAAATCAGCGAAAT-3’, N1 Reverse, 5’-TCTGGTTACTGCCAGTTGAATCTG-3’, N1 FAM probe, 5’-(FAM)ACCCCGCATTACGTTTGGTGGACC(3’-BHQ-1)-3’, RP Forward, 5’-AGATTTGGACCTGCGAGCG-3’, RP Reverse, 5’-GAGCGGCTGTCTCCACAAGT-3’, and RP Cy5 Probe 5’-(Cy5)TTCTGACCTGAAGGCTCTGCGCG(3’-BHQ-3)-3’. 20 µL reactions (15 µL master mix + 5 µL RNA) were prepared using TaqPath™ 1-Step Multiplex Master Mix (No ROX) (ThermoFisher Cat. # A28523) and 20X primer/probe mix. The final primer concentrations per reaction were: N1 Forward & Reverse Primers (400 nM), N1 Probe (200 nM), RF Forward & Reverse Primers (200 nM), and RP Probe (100 nM). The plate was run on the QuantStudio 7 under the following conditions: 25°C for 2 min, 53°C for 10 min, 95°C for 2 min, and 45 cycles of 95°C for 3 sec then 60°C for 30 sec. Fluorescence was detected at the end of each 60°C cycle.

### Library Preparation and Sequencing

Paragon Genomics’ CleanPlex® FLEX SARS-CoV-2 Panel (Cat. 918014) for Illumina platforms was used to prepare sequencing libraries (starting concentration of 10-50 ng RNA per sample). As a positive control, sequencing libraries were also prepped for VR-1986D, Genomic RNA from SARS-Related Coronavirus 2, Isolate USA-WA1/2020. Library quality and concentration was assessed via fragment analysis using Advanced Analytics’ High Sensitivity NGS Fragment Analysis Kit (Cat. DNF-474-0500). Library quality ratio scores (QRS) were determined by dividing the fragment analysis 250-350 bp peak concentration (ng/µL) by 50-190 bp peak concentration (ng/µL): excellent (QRS >10), Good (QRS 1.0 – 10), Fair (QRS <1 and >0.5), Poor (QRS < 0.5). Libraries with QRS > 1.0 were denatured and diluted to a final loading concentration of 10 pM following the Illumina MiSeq System Denature and Dilute Libraries Guide (Document # 15039740 v10), and then sequenced on the MiSeq system at 2 x 151 bp using the MiSeq v3, (600 cycle) kit (Illumina, Cat. MS-102-3003).

### Bioinformatics

Illumina adaptor sequences were trimmed using the BaseSpace Onsite FASTQ Toolkit v1.0.0. Adapter trimmed FASTQ files were aligned to the SARS-CoV-2 reference genome (NC_045512.2) using Illumina’s DRAGEN Bio-IT Platform. Primer sequences were removed using the Linux environment fgbio toolkit (bcftools) and a tab delimited file with primer genomic coordinates provided by Paragon Genomics. The DRAGEN was used to create variant call files from primer trimmed BAM files and consensus FASTA files were created using the fgbio toolkit. Genome coverage uniformity and mapping was visualized in IGV (BAM and VCF files).

## Data Availability

All data supporting the findings of this study are available from the corresponding author, TG, upon request. All requests are subject to PA-clearance.

## Disclaimer

The views expressed are those of the author and are not necessarily the official views of, or endorsed by, the U.S. Government, the Department of Defense, or the Department of the Air Force. No Federal endorsement of the platforms to produce this research data is intended. The voluntary, fully informed consent of the subjects used in this research was obtained as required by 32 CFR 219 and DoDI 3216.02_AFI 40-402.

## Acknowledgements

The authors would like to thank the laboratories who provided the excess clinical specimens used in this research.

## References

1. Li X, Wang W, Zhao X, Zai J, Zhao Q, Li Y, et al. Transmission dynamics and evolutionary history of 2019-nCoV. J Med Virol. 2020;92: 501–511. doi:10.1002/jmv.25701

2. Borges V, Alves MJ, Amicone M, Isidro J, Zé-Zé L, Duarte S, et al. Mutation rate of SARS-CoV-2 and emergence of mutators during experimental evolution. bioRxiv. 2021; 2021.05.19.444774. doi:10.1101/2021.05.19.444774

3. van Dorp L, Richard D, Tan CCS, Shaw LP, Acman M, Balloux F. No evidence for increased transmissibility from recurrent mutations in SARS-CoV-2. Nature Communications. 2020;11: 5986. doi:10.1038/s41467-020-19818-2

4. Long SW, Olsen RJ, Christensen PA, Subedi S, Olson R, Davis JJ, et al. Sequence Analysis of 20,453 Severe Acute Respiratory Syndrome Coronavirus 2 Genomes from the Houston Metropolitan Area Identifies the Emergence and Widespread Distribution of Multiple Isolates of All Major Variants of Concern. Am J Pathol. 2021;191: 983–992. doi:10.1016/j.ajpath.2021.03.004

5. Cyranoski D. Alarming COVID variants show vital role of genomic surveillance. Nature. 2021;589: 337–338. doi:10.1038/d41586-021-00065-4

6. Coughlin SS, Yiǧiter A, Xu H, Berman AE, Chen J. Early detection of change patterns in COVID-19 incidence and the implementation of public health policies: A multi-national study. Public Health Pract (Oxf). 2021;2: 100064. doi:10.1016/j.puhip.2020.100064

7. Tillett RL, Sevinsky JR, Hartley PD, Kerwin H, Crawford N, Gorzalski A, et al. Genomic evidence for reinfection with SARS-CoV-2: a case study. Lancet Infect Dis. 2021;21: 52– 58. doi:10.1016/S1473-3099(20)30764-7

8. National Center for Immunization and Respiratory Diseases (NCIRD), Division of Viral Diseases, CDC. SARS-CoV-2 Variant Classifications and Definitions. 2021 May. Available: https://www.cdc.gov/coronavirus/2019-ncov/variants/variant-info.html

9. Davies NG, Abbott S, Barnard RC, Jarvis CI, Kucharski AJ, Munday JD, et al. Estimated transmissibility and impact of SARS-CoV-2 lineage B.1.1.7 in England. Science. 2021;372. doi:10.1126/science.abg3055

10. Jangra S, Ye C, Rathnasinghe R, Stadlbauer D, Krammer F, Simon V, et al. The E484K mutation in the SARS-CoV-2 spike protein reduces but does not abolish neutralizing activity of human convalescent and post-vaccination sera. medRxiv. 2021. doi:10.1101/2021.01.26.21250543

11. Abu-Raddad LJ, Chemaitelly H, Butt AA. Effectiveness of the BNT162b2 Covid-19 Vaccine against the B.1.1.7 and B.1.351 Variants. N Engl J Med. 2021 [cited 29 May 2021]. doi:10.1056/NEJMc2104974

12. Shinde V, Bhikha S, Hoosain Z, Archary M, Bhorat Q, Fairlie L, et al. Efficacy of NVX-CoV2373 Covid-19 Vaccine against the B.1.351 Variant. N Engl J Med. 2021;384: 1899– 1909. doi:10.1056/NEJMoa2103055

13. Wang J, Kaperak C, Sato T, Sakuraba A. COVID-19 reinfection: a rapid systematic review of case reports and case series. J Investig Med. 2021. doi:10.1136/jim-2021-001853

14. Center for Devices and Radiological Health, FDA. In Vitro Diagnostics EUAs - Molecular Diagnostic Tests for SARS-CoV-2. 2021 May. Available: https://www.fda.gov/medical-devices/coronavirus-disease-2019-covid-19-emergency-use-authorizations-medical-devices/in-vitro-diagnostics-euas-molecular-diagnostic-tests-sars-cov-2.

15. Lippi G, Simundic A-M, Plebani M. Potential preanalytical and analytical vulnerabilities in the laboratory diagnosis of coronavirus disease 2019 (COVID-19). Clin Chem Lab Med. 2020;58: 1070–1076. doi:10.1515/cclm-2020-0285

16. Marcus JE, Frankel DN, Pawlak MT, Casey TM, Blackwell RS, Tran FV, et al. COVID-19 Monitoring and Response Among U.S. Air Force Basic Military Trainees - Texas, March-April 2020. MMWR Morb Mortal Wkly Rep. 2020;69: 685–688. doi:10.15585/mmwr.mm6922e2

17. Office of the Commissioner, FDA. Coronavirus (COVID-19) Update: FDA Authorizes First Antigen Test to Help in the Rapid Detection of the Virus That Causes COVID-19 in Patients. 2020 May. Available: https://www.fda.gov/news-events/press-announcements/coronavirus-covid-19-update-fda-authorizes-first-antigen-test-help-rapid-detection-virus-causes.

18. Centers for Disease Control and Prevention. Interim Guidance for Antigen Testing for SARS-CoV-2. 2021 May. Available: https://www.cdc.gov/coronavirus/2019-ncov/lab/resources/antigen-tests-guidelines.html

19. Joint Base San Antonio. MEDCoE Adds Antigen Testing Lab to COVID-19 Mitigation Strategy. 2021 Feb. Available: https://www.jbsa.mil/News/News/Article/2481284/medcoe-adds-antigen-testing-lab-to-covid-19-mitigation-strategy/.

20. Kumar M, Iyer SS. ASSURED-SQVM diagnostics for COVID-19: addressing the why, when, where, who, what and how of testing. Expert Rev Mol Diagn. 2021;21: 349–362. doi:10.1080/14737159.2021.1902311

21. Rasmi Y, Li X, Khan J, Ozer T, Choi JR. Emerging point-of-care biosensors for rapid diagnosis of COVID-19: current progress, challenges, and future prospects. Anal Bioanal Chem. 2021; 1–23. doi:10.1007/s00216-021-03377-6

22. Chiara M, D’Erchia AM, Gissi C, Manzari C, Parisi A, Resta N, et al. Next generation sequencing of SARS-CoV-2 genomes: challenges, applications and opportunities. Brief Bioinform. 2021;22: 616–630. doi:10.1093/bib/bbaa297

23. World Health Organization. Genomic sequencing of SARS-CoV-2: a guide to implementation for maximum impact on public health. WHO; 2021 Jan. Available: https://www.who.int/publications/i/item/9789240018440

24. Food and Drug Administration. BioFire COVID-19 Test: Instructions for Use under an Emergency Use Authorization (EUA) Only. 2020. Available: https://www.fda.gov/media/136353/download

25. Food and Drug Administration. BINAXNOW COVID-19 AG CARD (PN 195-000) – INSTRUCTIONS FOR USE. 2020. Available: https://www.fda.gov/media/141570/download

26. Ito Y. Clinical Diagnosis of Influenza. Methods in molecular biology. 2018;1836: 23–31.

27. Centers for Disease Control and Prevention. Interim Laboratory Biosafety Guidelines for Handling and Processing Specimens Associated with Coronavirus Disease 2019 (COVID-19). Available: https://www.cdc.gov/coronavirus/2019-ncov/lab/lab-biosafety-guidelines.html.

28. DHHS, CDC & NIH. Biosafety in Microbiological and Biomedical Laboratories, 6th edn. (US Government Printing Office, Washington, DC, June 2020). https://www.cdc.gov/labs/BMBL.html. Available: https://www.cdc.gov/labs/BMBL.html

29. Nazario-Toole AE, Xia H, Gibbons TF. Whole-genome Sequencing of SARS-CoV-2: Using Phylogeny and Structural Modeling to Contextualize Local Viral Evolution. Military Medicine. 2021 [cited 29 May 2021]. doi:10.1093/milmed/usab031

30. Research Use Only 2019-Novel Coronavirus (2019-nCoV) Real-time RT-PCR Primers and Probes. Available: https://www.cdc.gov/coronavirus/2019-ncov/lab/rt-pcr-panel-primer-probes.html

31. Kevadiya BD, Machhi J, Herskovitz J, Oleynikov MD, Blomberg WR, Bajwa N, et al. Diagnostics for SARS-CoV-2 infections. Nature Materials. 2021;20: 593–605. doi:10.1038/s41563-020-00906-z

